# Enhancing Two-Dimensional Control via Single-Channel Haptic Feedback: A Multi-dimensional Encoding Strategy

**DOI:** 10.1101/2024.12.09.24317386

**Authors:** T R Benigni, A Pena, S Kuntaegowdanahalli, J J Abbas, R Jung

## Abstract

Recently a multidimensional encoding approach to direct stimulation of a single site in the median nerve showed that an intensity and flutter frequency could be perceived and graded. It is unclear whether these two dimensions are able to convey useful information to participants performing control tasks or whether they improve on typical intensity only modulation approach. Eleven participants performed experiments to assess the efficacy of multidimensional stimulation. In a set of discrete matching tasks, participants were able to correctly identify all thirteen discrete stimulation points better than chance. In a modified center-out task, and three separate extremes-in tasks, all seventeen changes in the stimulation parameters could be detected better than chance. Performance of the discrete task using a multidimensional approach showed increased information transfer compared to the individual modulation of intensity or flutter frequency. The results suggest that multidimensional encoding is a promising approach for increasing information throughput in sensory feedback systems. By investigating a multimodal encoding approach, this study offers valuable insights into haptic feedback through peripheral nerve stimulation. This haptic feedback might offer pronounced benefits for virtual reality applications and individuals with upper limb amputations opening avenues for an enhanced sensory feedback experience.

## Introduction

Haptic feedback using various stimulation modes including vibration and electrostimulation has improved performance in teleoperation, created more immersive virtual reality environments, and improved clinical outcomes for users of myoelectric prostheses. Just because a stimulation approach provides a graded percept does not mean it will perform well as a haptic feedback system. Lag times between stimulation and perception and ambiguity between graded percepts leading to low information transfer could affect the feasibility of a given approach to providing meaningful haptic feedback. The same might be true of any novel stimulation approach, and each approach must be assessed as the inclusion of haptics becomes more widespread.

In a laboratory setting, the usefulness of haptic feedback is typically assessed by comparing task performance with and without feedback. Object discrimination tasks have been used to find the usefulness of this feedback to people with amputations and in virtual reality environments. Matching tasks have been used to assess vibrotactile feedback’s ability to deliver graded levels of force percepts and assessing the amount of information conveyed by different placements of vibrotactors for both prosthetic use and teleoperation. Center-out tasks have also been used to assess the usefulness of haptic feedback by measuring how well stimulation from vibrotactors, or brain-computer interfaces can be used to detect changes in the direction of stimulation.

Recently, a multidimensional encoding approach for haptic feedback provided by electrically stimulating the peripheral nerves has been developed. Haptic feedback through direct stimulation of the nerves causes perceived sensations that are distally referred or perceived in an area distal to the stimulation site. This form of feedback is also somatotopically matched, for example, stimulation of the ulnar nerve can elicit sensations perceived on the phantom ring and little finger of a person with an amputated limb. In all these studies, graded intensity of haptic percepts was modulated by changing stimulation pulse parameters such as pulse width, pulse amplitude, or pulse frequency within a comfortable range [20]. By using this multidimensional encoding approach, percepts of frequency in the flutter range (5-25 Hz), and graded intensity could be delivered simultaneously and perceived in a single distally referred area of the hand. Specifically, the multidimensional encoding was done by providing repeated bursts of stimulation pulses. The percept flutter frequency was modulated by altering the interburst interval while the percept intensity was modulated by changing the pulses sent within the burst following a charge rate model.

Multiple studies have shown that the ability to convey multiple percepts can improve performance in functional tasks and increase the information transferred especially when the different percepts can be independently graded and differentiated. Since the multidimensional encoding approach can convey both the percepts of flutter frequency and percept intensity and offer graded information about them, we hypothesized that a multidimensional encoding approach, altering flutter frequencies through BP modulation, and intensity through charge rate modulation can be used to perform two-dimensional control tasks.

To this end, in this study participants performed a series of control tasks while receiving stimulation inside a two-dimensional stimulation parameter space, where the x-direction corresponded to increasing flutter frequency and the y-direction to increasing intensity. The ability to recognize different points in the stimulation space was assessed through a discrete matching task, in which participants had to match the stimulation they received to a discrete point in the parameter space. We determined if participants could differentiate between stimulation trains composed of different combinations of frequency and intensity better than chance. To assess whether participants could track simultaneous changes in the stimulation parameters, we assessed whether participants could track changes in the stimulation levels significantly better than chance. A modified center-out task was used to assess this. Others have seen that decreasing intensity causes increased difficulty in detecting changes in both intensity and frequencies. We tested whether this amplitude dropout is true by comparing the performance of multiple change-in-direction tasks which began at the limits of the intensity and frequency range. The multidimensional encoding approach would be a benefit only if it improves upon the current modulation of intensity only stimulation approach. We therefore determined if it provided increased information transfer compared to the conventional intensity modulation encoding approach. This was tested by comparing the information transfer delivered by the multidimensional, and conventional approach during the discrete matching tasks. The results of this study show that participants can accurately use this multidimensional encoding approach as a source of feedback when performing tasks that involve differentiation and tracking. Hence, this approach might improve conventional methods of providing graded percepts, creating more realistic tactile percepts for haptic feedback.

## Results

### Sensory characterization

Eleven participants (n = 11) received stimulation of their median nerve through non-invasive stimulating electrodes placed on the wrist. The pulse amplitude (PA), pulse width (PW), pulse frequency (PF) within a stimulation burst, the interburst interval (IBI), and the burst period (time between initiation of two consecutive busts) were changed. Each participant reported receiving comfortable distal sensations in the area innervated by the median nerve. Local sensations under the electrodes were not reported once calibration was completed. After calibration, an average stimulation pulse amplitude of 1996 ± 306 µA, a comfortable PW range of 469– 699 µs, and fusion to saturation PF range of 62 – 180 Hz were found. Participants performed the entire three-hour session without having to readjust calibration parameters.

Using the multidimensional encoding approach, all participants were able to perceive the two simultaneous percepts of intensity and flutter frequency. Intensity was modulated following the charge rate model by changing the PW and PF of the stimulation pulses inside of 40ms bursts. The flutter frequency was then modulated by changing the off time or IBI between each burst between 10 – 160 ms.

### Participants could identify multiple points in the stimulation space better than chance

In a discrete matching task participants matched the intensity, flutter frequency, or both dimensions of the stimulation they received with a target in the stimulation parameter space as shown in Figure 5a. Three iterations of the task were performed. In the first iteration, only the flutter frequency of stimulation was modulated and participants had to match the stimulation to four targets on the x-axis of the stimulation parameter space. For the second iteration only the intensity of the percept was modulated and participants had to match the stimulation to four targets on the y-axis of the stimulation parameter space. In the third set, both the intensity and frequency of the targets were changed, and participants matched the stimulation to five targets distributed across the stimulation parameter space.

Chi-squared analysis of all three discrete tasks showed that participants could match all discrete stimulation targets provided to them better than chance (p < 0.0001 per task) as seen in Figure 1a. During the task where only the intensity of the stimulation was modulated, participants performed with an average correct percentage of 95% ± 12.5%, 66% ± 13.3%, 60% ± 28.2%, and 77% ± 24% for intensity values of 20%, 40%, 60%, and 80% of the QR range, respectively. T-tests of each intensity target compared to chance showed significantly better performance with the highest p-value at p < 0.003. In the task where only the flutter frequency was modulated, an average correct percentage of 68% ± 33%, 66% ± 13.3%, 53% ± 25%, and 88% ± 9% for intensity values of 20%, 40%, 60%, and 80% of the burst period range respectively were obtained. The highest p-value was (p = 0.008). As illustrated in Figure 1b, in the discrete matching task where both parameters were modulated, participants successfully matched the stimulation parameters with an average correct percentage per target of 89% ± 9%, 89% ± 9%, 73% ± 24%, 91% ± 10%, 91% ± 10% ((20%, 20%), (20%, 80%),(50%, 50%), (80%, 20%), (80%, 80%) for the BP and QR range respectively). In the tasks where both parameters are modulated, the highest p -value recorded was (p < 0.0001).

**Figure 1.**
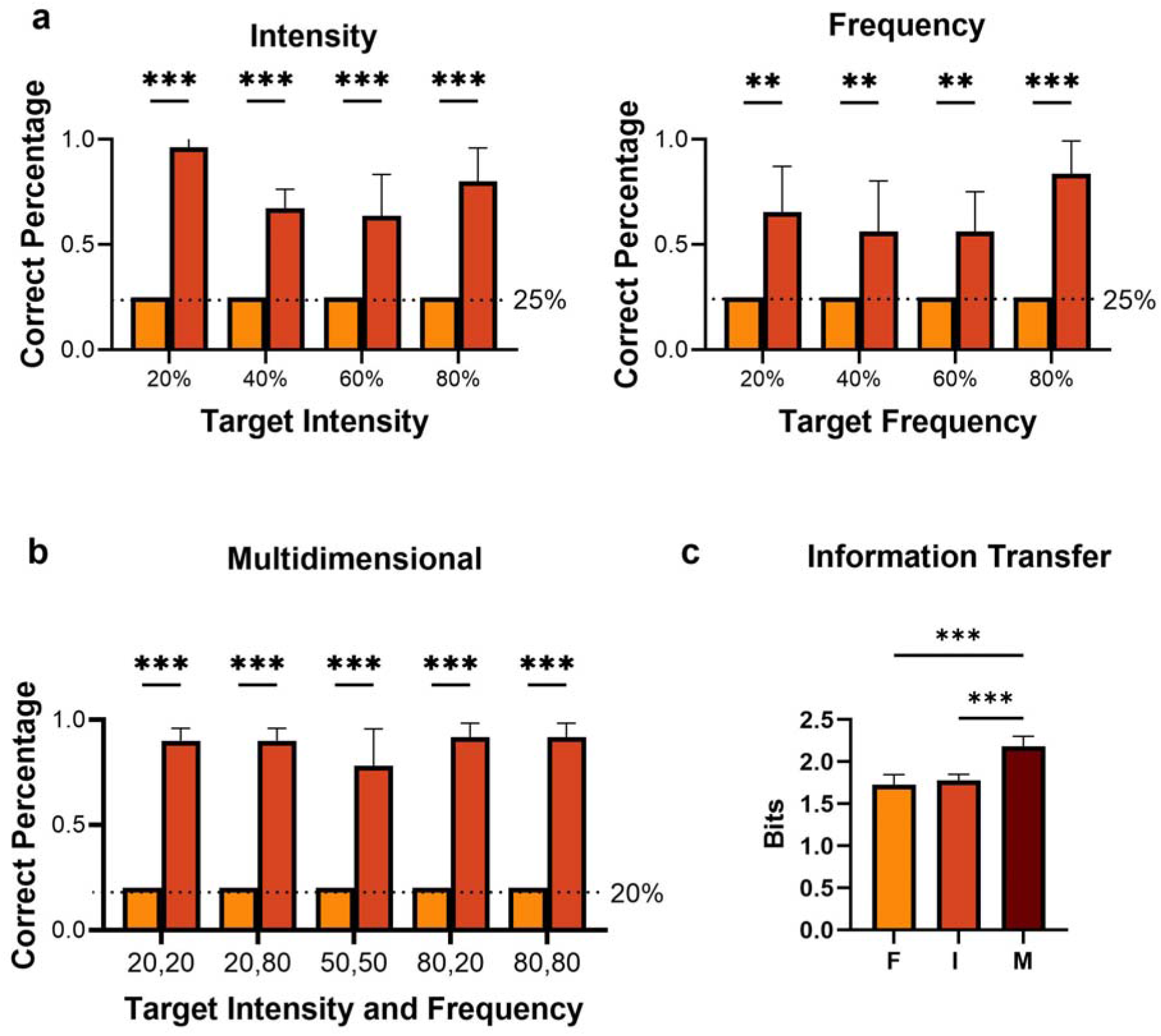
Detection of discrete stimulation targets. (**a**) Group average (n=11) correct response percentage for each target in the discrete control task where only the intensity or flutter frequency were modulated. On the X-axis, the target’s stimulation value is labeled. Each target’s average response rate is compared to the chance response rate (25%). Asterisks indicate the results of a T-test. *** = *p* < 0.001. (**b**) Group average correct response rate for the multidimensional discrete task. In this task, both the intensity and frequency of each target were modulated. The target’s stimulation value is labeled on the X-axis. Each target’s average response rate is compared to the chance response rate (20%). Asterisks indicate results of a T-test. ** = *p* < 0.01, *** = *p* < 0.001 (**c**) The information transfer rate for each of the discrete tasks. On the X-axis is the name of the discrete task assessed: frequency (F), intensity (I), and multidimensional (M). An ANOVA was used to discover significant differences between the groups indicated by the asterisks. *** = *p* < 0.001

### Participants could detect simultaneous changes in intensity and frequency better than chance

A modified center-out task which we termed a change-in-direction task was used to assess how well participants could detect changes in the stimulation parameters. When performing the change-in-direction task, participants were able to tell the difference between changing flutter frequency percepts with an average correct response rate of 97% ± 0.06% in either an increasing or decreasing direction (*p* < 0.0001). Increases in intensity were detected with 100% accuracy after training, while decreases were detected with 91% ± 19% accuracy. As seen in Figure 2a when both intensity and flutter frequency were changed simultaneously, participants were still able to detect changes in the stimulation better than chance, with an average correct percentage of 68% ± 27%, 87% ± 19%, 82% ± 17%, and 89% ± 25% for the (−x, −y), (−x, +y), (+x, −y), (+x, +y) changes, respectively. All participants could detect the changes in intensity and flutter frequency better than chance (Figure 2b).

**Figure 2.**
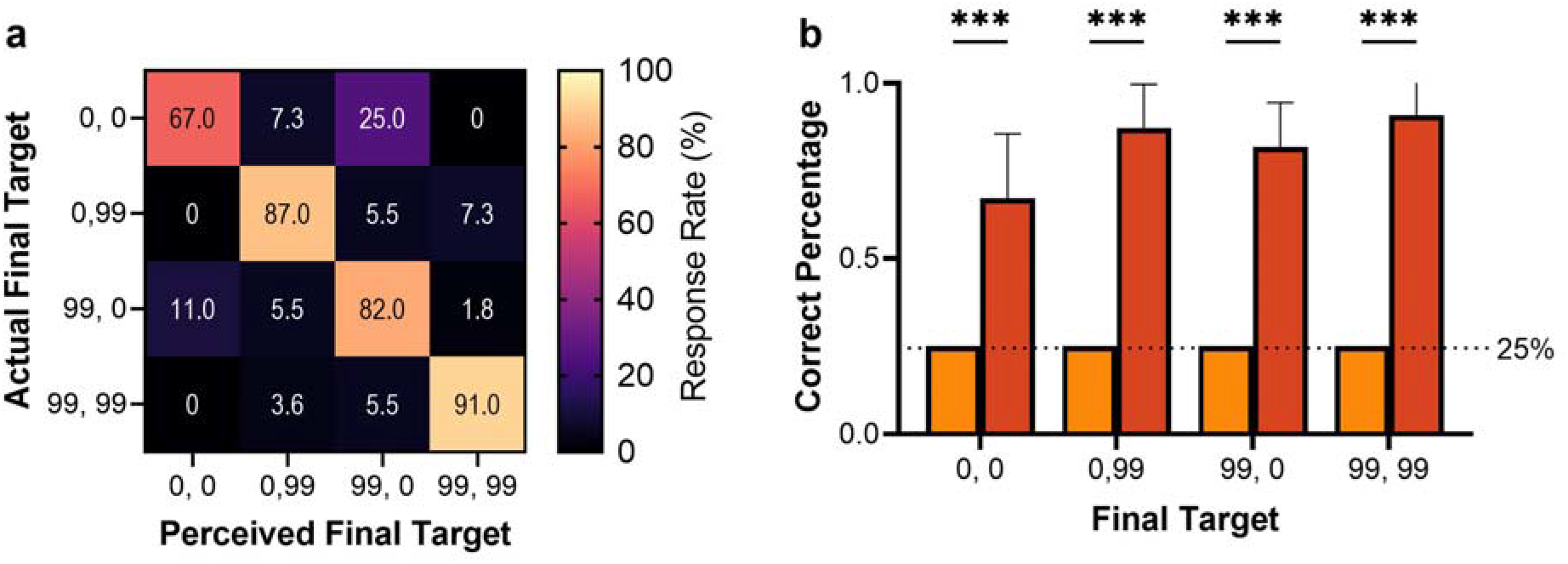
Detection of simultaneous frequency and intensity changes. (a) Confusion matrix quantifying overall performance in a center-out task. During the center-out task, each stimulation train starts at 50% of both the frequency and intensity ranges and moves to the maximums or minimums of flutter frequencies and intensity simultaneously. The Y-axis of the matrix shows the final target to which the stimulation went. The X-axis shows where the participants perceived the stimulation went, or the answer given by the participant. Each block represents the frequency of responses provided by all participants when they were presented with a stimulation train (actual) and classified (perceived). (b) The average correct (n = 11) response percentage for each final target compared to chance (25% correct). T-tests or Mann-Whitney tests were used to determine whether there was a significant difference compared to chance (*** = *p* < 0.001)

### Multidimensional encoding had significantly greater information transfer than intensity or flutter frequency modulation

The information transfer on stimulation using a multidimensional encoding approach was significantly higher than when using intensity or flutter frequency modulation alone as seen in Figure 1c. Intensity modulation alone transmitted an average of 1.76 bits ± 0.05, flutter frequency modulation transmitted 1.74 bits ± 0.1, and the multidimensional modulation conveyed 2.16 bits ± 0.11. Results of an ANOVA comparing the three encoding approaches called for a post hoc test (*F* = 61.37, *p* < 0.001). The results of a Tukey’s post hoc analysis showed that the information transfer with a multidimensional encoding approach is significantly greater than both intensity and flutter frequency modulation alone *p* < for both comparisons.

### Detecting changes in the stimulation was significantly harder when starting at a high intensity and slow flutter

In another task, participants were asked to track changes in the stimulation starting from the extremes of the stimulation parameters and following a preset course. Each iteration of the task was titled with the location of the starting parameters in the stimulation parameter space as seen in Figure 5c. The first iteration of the task started from the lowest intensity and slowest frequency (Bottom-Left). the second iteration corresponded to the highest intensity but the slowest flutter frequency (Top-Left), and the third iteration corresponded to the fastest flutter frequency and lowest intensity (Bottom-Right). Each iteration was presented in random order.

Participants were able to detect changes in stimulation in all three iterations of the extremes-in task better than chance regardless of the starting position as seen in Figure **3**a; (χ^2^= 68.31, *p* < 0.0001) for the Bottom-Left task, (χ^2^= 82.73, *p* < 0.0001) for the Bottom-Right task, and (χ ^2^= 68.31, *p* < 0.0001) for the Top-Left task. ANOVA showed a significant difference in performance in the three extremes-in tasks (*F=* 7.95, *p* = 0.02). A Tukey’s post hoc analysis showed that performance in the Top-Left extremes-in task was significantly worse than the Bottom-Left extremes-in task (*p* = 0.01) and the Bottom-Right extremes-in task *p* = 0.002 as seen in Figure **3**b. These results reflect qualitative responses obtained through a verbal survey on task preference during the experimental sessions. Six out of the eleven participants reported the Bottom-Right extremes-in as their preferred task, five out of the eleven preferred the Bottom-Left. No participants indicated that the Top-Left extremes-in task was their preferred task.

## Discussion

This work assessed if a multidimensional encoding approach could convey task-specific haptic feedback through control tasks. The results support our hypothesis that a multidimensional encoding approach, modulating flutter frequencies through BP modulation, and intensity through QR modulation can be used to perform two-dimensional control tasks. The participants could differentiate discrete stimulation parameters and changes in multidimensional stimulation. They could perform all tasks provided to them with performances significantly better than chance. All participants could differentiate between the thirteen discrete points in the stimulation parameter space, and participants could detect changes in seventeen different directions through the parameter space better than chance. These changes in stimulation and the discrete points were not presented in a single trial. Instead, the targets were spread out over different tasks and trials. A single trial having 13 different points would have been extremely difficult for anyone as increasing the possible choices beyond four or five options is known to increase difficulty.

The multidimensional encoding approach increases the information transmitted through a single stimulation channel. This encoding approach has more information than intensity modulation alone. The information transfer of the intensity encoding matches values found in the literature when stimulating the peripheral nerve through microneurography. Results from a few case studies on interneural approaches to providing PNS suggest similar information transfer in their conveyed signals. With the addition of perceived frequency to the encoding approach, the information transfer exceeds earlier limits of 1.5 - 2 bits in unimodal haptic feedback. This shows that the information transfer of a multidimensional encoding approach is greater in participants with intact peripheral nerves.

While participants could still detect changes in the stimulation beginning from a high intensity/low frequency, they performed significantly worse when the stimulation started at a low intensity/low frequency or low intensity/high frequency. Thus, detecting changes in both intensity and frequency became more difficult when the intensity was near its maximum. A phenomenon known as amplitude dropout, as experienced in other studies [31,38], could explain this reduction in performance. As these studies described, the simple drop in the intensity of the stimulation makes it difficult to differentiate any other feature or even the degree of drop in intensity. During the Top-Left extremes-in task trials where only the burst period changed, participants successfully identified the change with an 85% correct response rate. In the two trials where the intensity decreased, the correct response rate dropped to either 47% or 53%, respectively, as shown in Figure 3a. The results of extremes-in task could be used to inform how to convey two separate streams of information, such as two changing sensor values on a prosthetic or from a VR environment. Our results suggest that sensor values should be mapped such that the stimulation starts at a low intensity, when detecting a low grasp force or hand aperture values, etc. However, our results also suggest that sensor values could be mapped such that the stimulation starts at either a high or low flutter frequency, when detecting low sensor values, without affecting the ability to detect changes.

**Figure 3.**
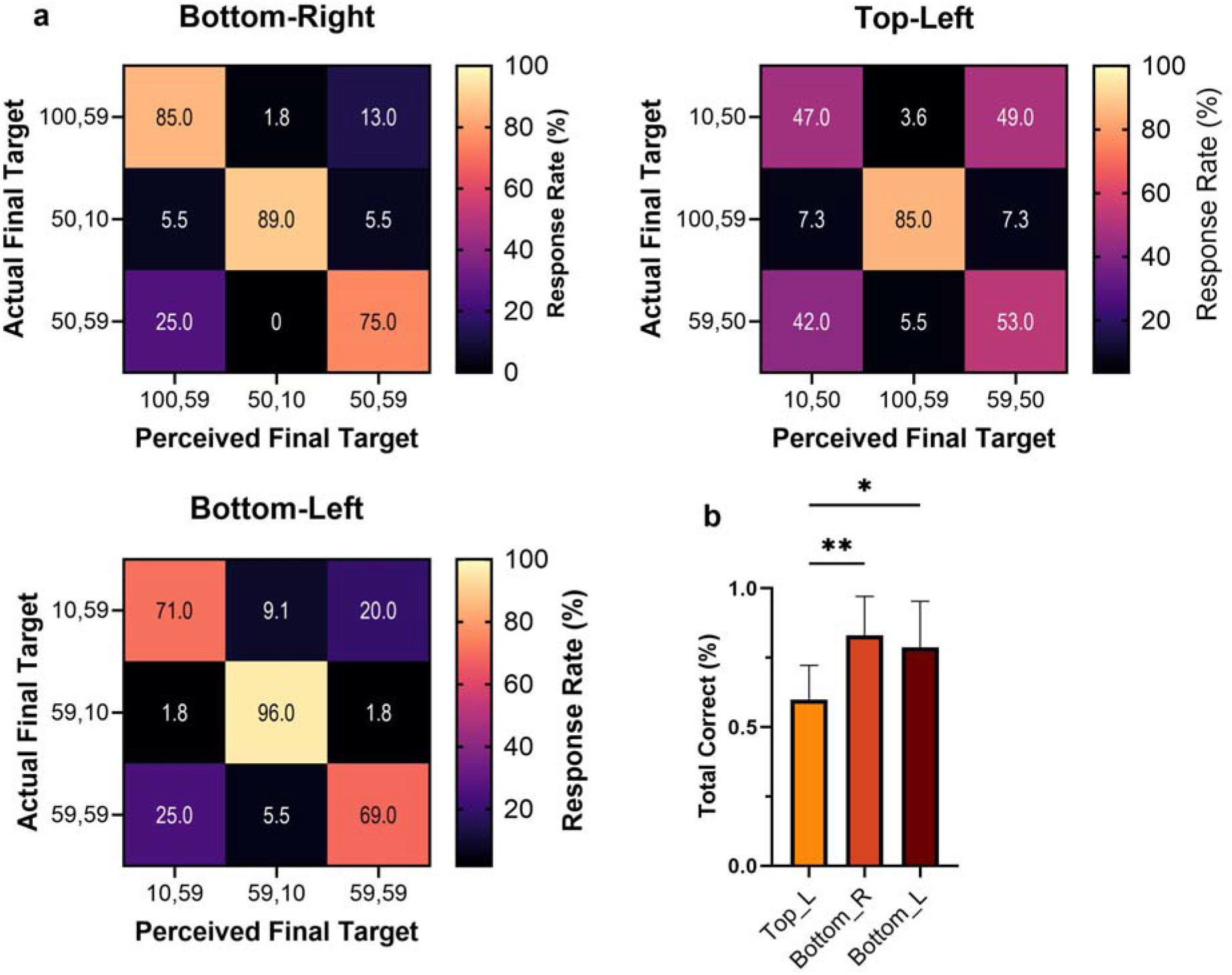
Participants could detect changes at the intensity and frequency range limits. Three confusion matrices for each corner in the extremes-in tasks. The title of each confusion matrix indicates the origin point for each task. The final frequency and intensity target in the extremes-in task is on the Y-axis. The X-axis shows the perceived final intensity and frequency target. Each block represents the frequency of responses provided by all participants when they were presented with a stimulation train (actual) and classified (perceived). (**b**) Group average (n = 11) correct response percentage of the three tested extremes with SD. An ANOVA and subsequent Tukey’s were conducted to find any significant difference between tasks (*= *p* < 0.05, ** = *p* < 0.01)

Although all the participants were given ample training to perform the discrete matching task, when performing the data collection phase of the discrete matching task, participants were not given any visual feedback about where in the stimulation parameter space they were supposed to move the joystick. This may have led to a few errors caused by inaccuracy in controlling the joystick rather than misclassifying the target. The error caused by the lack of visual feedback was systematic, as all the matching tasks were performed in the same manner, just with different combinations for flutter frequency and intensity. However, the lack of visual feedback added a degree of difficulty to the matching tasks which may not have been necessary.

Participants use many internal and external sources of information that may influence their responses while performing tasks. This phenomenon is known as response bias Our study did not account for possible bias while performing the tasks provided. We accounted for randomization by making all the presentations for every task double blinded [38]. We also accounted for the matching bias by adding a 0.5 second delay between the end of a stimulation and the ability to record a response [39]. What we did not account for was participants “counting” the number of the presentations and using that to improve their performance [40]. To assess this, a secondary analysis consisting of a set of paired T-tests was performed.. The performance between the first and last five targets presented in the matching and center-out tasks was compared; *p =* 0.32 for the intensity discrete matching task, *p =* 0.19 for the flutter frequency discrete matching task, *p =* 0.62 for the multidimensional discrete matching task, and *p =* 0.31 for the center-out task This indicates that there was no significant difference between the performance at the beginning and end of each task indicating that although we did not control for “counting” it did not occur.

Participants performed the tasks provided in a single session and had never received peripheral nerve stimulation for haptic feedback before the session began. This lack of familiarity with the multidimensional encoding approach may have led to reduced performance. The training provided during the session allowed them to reach an intra-session performance plateau for every task, ensuring that the participants understood the task and had time to formulate a mental strategy. This intra-session plateau does not represent the best possible performance. Many others have suggested that participants’ performance would improve over a more extended training period [43-45]. Even though their performance was already better than chance at all the tasks presented, one of the participants said, “With experience, this would be functional.” This statement may reflect intuitiveness to the multidimensional encoding approach. It could indicate that haptic feedback using multidimensional encoding could be easily adopted during active daily use.

All participants were from a younger cohort. Age may have had a positive impact on our study as performance on tasks that use input devices such as joysticks have a negative correlation with age. However, an older cohort should also be able to use this multimodal encoding approach. Age has been shown to affect motor learning times, but not necessarily the transfer of the learning to new tasks. The latter suggests that an older population could likely learn to use this encoding approach, but possibly over multiple sessions. The current study did not investigate the fullness of learning that could have taken place, as we did not assess how well participants could perform novel tasks after an initial learning period, and these tasks were not performed longitudinally [48,49].

In other studies which included a matching task, participants used their contralateral hand to press on a force sensor to match force values conveyed through either neural stimulation or vibrotactile stimulation. In this study, participants moved a joystick to match specific points in the stimulation parameter space. As we intended to use this matching task to assess the differences in the information transfer between intensity, flutter frequency, and a multidimensional approach, a single method across all three encoding approaches seemed appropriate. This method of performing the matching task may lead to ambiguity in specific applications of this encoding approach.

In this study sensory feedback was provided by electrically stimulating the peripheral nerves of participants to elicit distally referred sensations. This method was chosen as it can elicit sensations on the phantom hands of people with an upper limb amputation. Providing these sensations on the phantom hand not only improves embodiment and acceptance of the prosthesis [6,50,51], but also improves performance in functional tasks [18,52] [37,53]. In addressing the functional benefits of sensory feedback, we demonstrated that our mulitmodal encoding approach increases the information transferred to participants with intact peripheral nerves. Future experiments are needed to assess whether this remains true for people with an upper limb amputation whose PNS has sustained an injury. Although transcutaneous stimulation of intensity percepts through the median nerve of people with an amputation has been conducted, the information transfer in those experiments was not published [37,53].

In summary, this work showed that a multidimensional encoding approach might be a feasible method of conveying haptic feedback. Results from this study show, for the first time, that participants could detect multiple different flutter frequencies and intensity values and changes to both flutter frequency and intensity with a greater information transfer than conventional approaches. This indicates that this encoding approach could be a suitable method for conveying enhanced haptic feedback. These experiments show that intensity and flutter frequency, when linearly mapped to an x and y axis, can be understood as two perpendicular dimensions. The ability to relate the percepts to two independent values implies that this encoding approach could help provide a participant with improved peripheral nerve based haptic feedback. Performance in actual functional tasks, such as an object discrimination task with a myoelectric prosthesis or a virtual box and blocks task, would still need to be assessed before concluding if this encoding approach has functional benefits in conveying haptic feedback.

## Methods

### Able-bodied Human Subjects

Six male and five female participants, with an average age of 25.2 ± 7.5 years were recruited to take part in this study. Ethical approval for the study protocol was obtained through the University of Arkansas Institutional Review Board (IRB # 2201379281). Participants were recruited for a single three-hour data collection session. Informed consent was obtained to publish the information/image(s) in an online open access publication.

### Experiment Setup

A multi-channel bio-stimulator (TDT IZ2-16H, Tucker-Davis Technologies, Alachua, FL USA) delivered charge-balanced, current-controlled biphasic, cathodic-first, rectangular pulses. To avoid local discomfort around the stimulating electrodes, a channel-hopping interleaved pulse scheduling (CHIPS) strategy was employed. The median nerve was targeted transcutaneously via four small, self-adhesive gel electrodes, two 15 mm by 20 mm stimulating electrodes and two 20 mm by 25 mm return electrodes (Rhythmlink International LLC, Columbia, SC, #STCUL15026, and STCUS25026,) placed around the left wrist. The stimulating electrodes were placed on the ventral aspect of the wrist approximately 3 cm from the distal radial crease, while the return electrodes were on the dorsal side. A small amount of current was sent through the electrodes to ensure they provided a distally referred sensation in the areas of the hand innervated by the median nerve. Low current (< 2500 µA for 500 µs) pulses were sent at 5 Hz while the participant reported if and where they perceived a sensation to ensure the electrode location elicited distally referred sensations.

Participants sat at a table with a display monitor placed in front of them to convey instructions, with their left arm on a cushioned support pad, with their medial side in contact with the pad. Depending on the task, they used their right hand to manipulate a control knob during the parameter calibration or a custom joystick during the experimental tasks as seen in Figure 4b. Experimental control software, written in Python 3 (version 3.11.3), managed the organization and execution of the experiments while storing the collected data. A custom stimulation control module was developed on the Synapse Software (version 96, Tucker-Davis Technologies (TDT), Alachua, FL USA) and controlled the stimulator.

**Figure 4.**
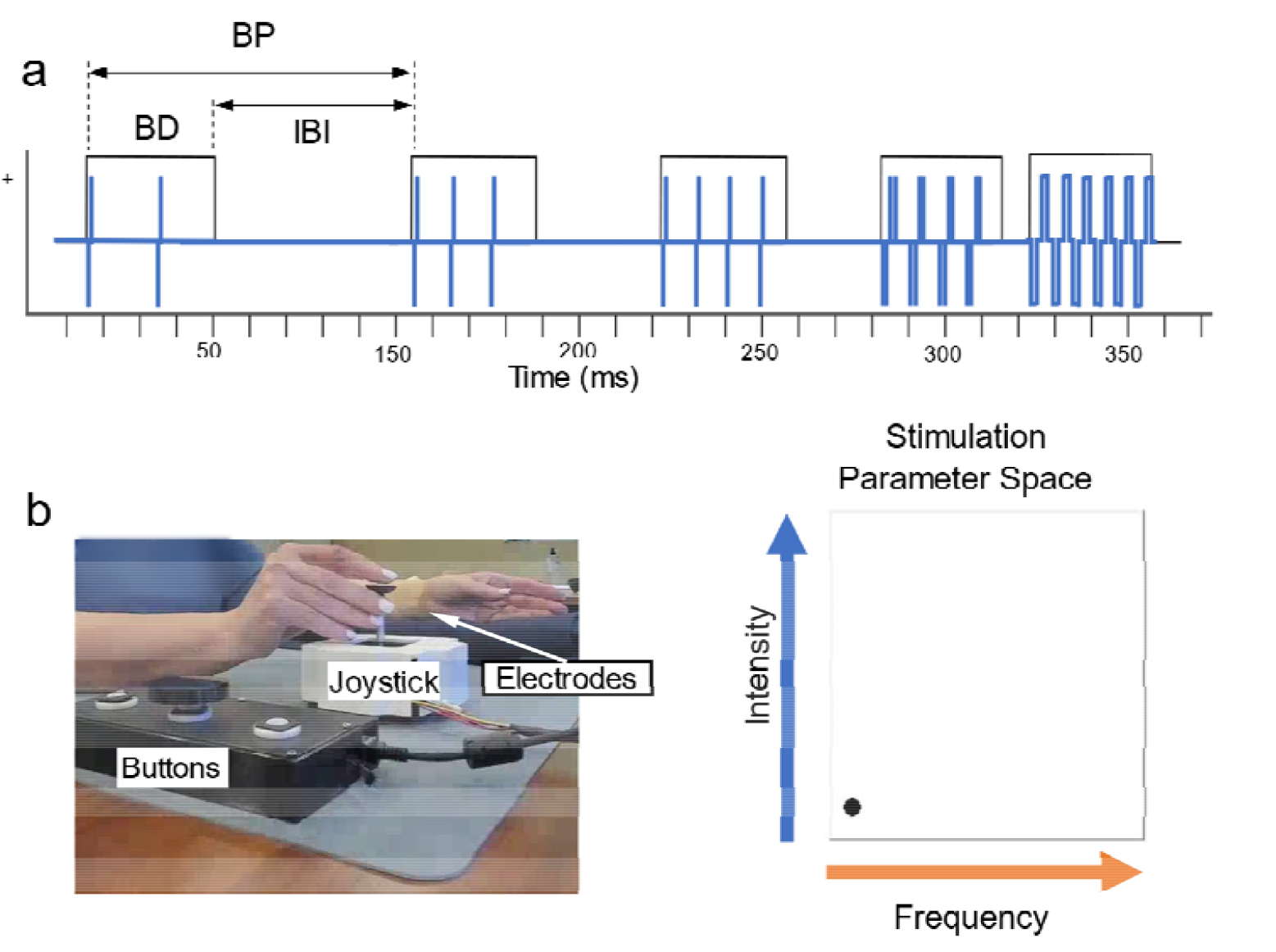
Multidimensional encoding approach and experimental setup during data collection. (**a**) A stimulation burst encoding approach was used to convey a percept of flutter frequency and intensity. The flutter frequency was modulated by changing the burst period (BP), given by the burst duration (BD) plus the interburst interval (IBI). BD remained constant while the IBI was varied. The intensity of the percept was modulated by changing the number and duration of biphasic, cathode-first rectangular pulses within each burst following the charge rate model. (**b**) Electrodes are placed on the left arm which rests on a pad. The right hand is used to manipulate a joystick to move a cursor inside a stimulation parameter space during the performance of the discrete control, center-out, and extremes-in tasks. The X-axis of the parameter space is linearly mapped to increasing flutter frequency, while the Y-axis is linearly mapped to increasing percept intensity.

### Encoding approach

As illustrated in Figure 4a, periodic bursts of stimulation pulses were provided with each burst period consisting of a constant burst duration (BD) of 40 ms, constant interphase gap of 100 µs, and an inter-burst interval (IBI) modulated between 10-160 ms to convey flutter frequencies between 5-20 Hz. A charge-rate (QR) encoding approach for intensity modulation was used to provide graded intensity percepts [21,54]. The intraburst pulse frequency (PF) and the pulse width (PW) were modulated along their comfortable ranges while the pulse amplitude (PA) was kept constant.

### Calibration

A participant-controlled calibration routine was used to find comfortable suprathreshold sensations. This approach was described in detail in other studies [11,54]. A strength-duration (SD) curve was generated to obtain the threshold of perceivable sensations. PA thresholds were collected for five different PWs between 200 -700 µs with a one hundred µs step at a low PF (5 Hz). Participants were given control of a knob the PA delivered when rotated clockwise. Thresholds were defined as when participants just begin to perceive a sensation. Each test PW was presented at least twice in random order. Responses were fitted to the Lapicque-Weiss’s model to obtain the SD curve. From each participants’ SD curve, a stimulation PA above the linear region of the curve was selected for the rest of the study. Typically, the PA was set at ∼25% above the threshold at a PW of 500 µs. To find the PW limits participants rotated the knob, which modulated PW between a range of 100-800 µs. For the lower limit of the PW, participants were instructed to find the PW where they reliably perceived a sensation; for the upper limit, they were asked to stay below a PW that led to an uncomfortable percept. For the PF limits, participants found the lowest possible frequency that was not perceived as pulsating, and then the highest frequency level at which the perceived stimulation intensity did not change.

### Stimulation parameter space

Each experimental task was represented in a two-dimensional stimulation space referred to as the stimulation parameter space as seen in Figure 4b. The X direction was mapped inversely to the range of the BP and therefore linear to the flutter frequency. The Y direction was mapped linearly to QR. Participants were given control of the joystick which moved a cursor on the screen in front of them. They were told that moving the cursor in the X direction modulated the frequency of the perceived sensation while the Y direction modulated its intensity. An exploratory training phase was used to introduce the two perceived dimensions. Using the joystick, participants freely explored different points along the stimulation parameter space. Once the exploratory training was over, participants were instructed that the joystick no longer controlled the stimulation but was to be used to respond to the control tasks they would be given

### Training

For each novel task, the participants first performed a target matching task with visual feedback about the stimulation parameters represented by a target on the screen in front of them as seen in Figure 5. This was followed by a training phase with delayed visual feedback. During the training phase, the participant performed a trial of the task without any visual feedback. On task completion, the target appeared along with the final position of the joystick cursor. Training tasks were shorter than the experimental version, having three random repetitions of all the targets in a specific task. Training continued until participants performed the task without errors or until a plateau of performance was reached. We defined a plateau of performance as three training sessions in which the number of correct responses remained constant (± 1 correct

**Figure 5.**
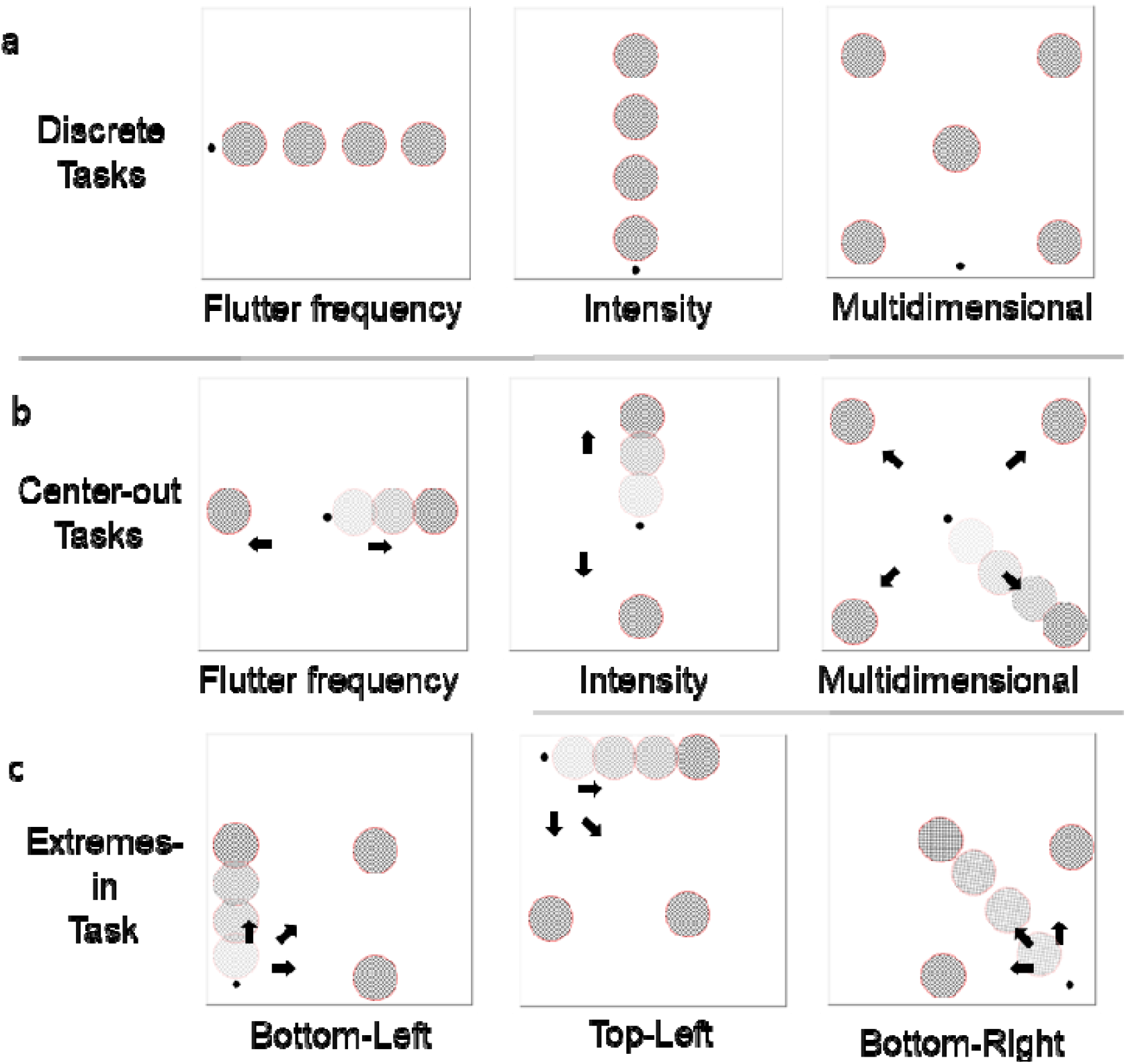
Visual display of the nine experimental tasks. Nine experimental tasks were performed in this study. The cursor’s position (small dot) shows the initial point where participants were instructed to start for each specific task. (**a**) In the discrete tasks, a 1.5-second stimulation train of a constant parameter equivalent to the target’s position was given to the participant. Participants were instructed to move the cursor to one of the possible targets. In (**b**) the center-out and (**c**) extremes-in tasks, the stimulation parameters changed at a constant rate over the course of three and a half seconds from the origin point to a final set of target parameters. Once the stimulation had stopped, participants moved the cursor to one of the final targets. In (**a**) and (**b**) Three versions of the task were performed, in two versions, only the intensity or flutter frequency of the target was modulated. In the third version, the target’s intensity and frequency was modulated multidimensionally. In (**c**) Bottom-left, Top-Left and Bottom-Right refer to the initial values used during that version of the extremes-in task.

### Discrete Control Task

To assess the ability to find combinations of stimulation parameters and differences in the information transfer, participants were asked to match the stimulation they received with a target in the stimulation parameter space Figure 5a, At the beginning of each trial, participants placed the cursor at the starting position, which varied depending on the version of the task. They then received a 1.5-second stimulation pulse train corresponding to a target in the parameter space. Once the stimulation ended, they were instructed to move the joystick to where they believed the target was and to press a button showing they had reached it. The response was correct if it was within a tolerance of ± 10% of the target value. The number of times a target was correctly identified was recorded as a correct percentage. From the performance of each participant, the information transfer was then determined. Information transfer can be defined as how well the information contained in a message can be used to perform certain tasks. This measure of performance reflects the integration of the sensorimotor system when processing sensory information. Information transfer reflects the effect that variables such as memory and signal clarity can have on task performance and is recorded in bits [9,56,57].

Three versions of the discrete matching task were conducted as seen in Figure 5a. In the first version, only the flutter frequency of stimulation was modulated. Participants had to correctly match four targets. These targets were set at 20%, 40%, 60%, and 80% of the BP range. Each target had a QR set at 50% of the QR range. In the second version only the intensity of the target was modulated. These targets were set at 20%, 40%, 60%, and 80% of the QR range. For the second version, the BP was kept at a constant 50% of its comfortable range. In the third set, both the intensity and frequency of the targets were changed. This set had five targets set at (20%, 20%), (20%, 80%), (50%, 50%), (80%, 20%), (80%, 80%) for the BP and QR ranges, respectively.

### Center-out Control Task

To assess the ability to perceive changes in the stimulation, participants were asked to do a variation of the center-out task [15,58]. At the beginning of each trial, the joystick and the cursor were placed in the center of the stimulation parameter space Figure 5b. The stimulation train began at 50% of the QR range and 50% of the BP range. Then, the BP (x) and QR (y) changed in one of eight directions. These directions included all combinations of ± x, ± y which correspond to a change of ± 30% in the QR and BP. Changes occurred over 1.5 seconds at a constant rate. Participants moved the joystick along one of the eight directions and pressed a button at the final position. Each stimulation train lasted 3.5 seconds. The train began at the initial stimulation value for one second. Then, over the next 1.5 seconds, the stimulation changed at a constant rate until it reached the final stimulation target and remained there for an additional second.

The center-out task was separated into three separate versions. In the first version, only the intensity of the stimulation changed in the -y, +y direction. The second version also had two targets with only the flutter frequency changing in the -x, +x directions. This was done to help participants learn to detect changes in the specific dimensions. In the last version of the center-out task, both the flutter frequency and intensity of the target changed. The targets for this trial changed in the following directions (−x, +y), (+x, +y), (−x, -y), (−x, +y). The number of times a target was correctly identified was recorded as a correct percentage.

### Extremes-in Control Task

To find if intensity drop affects how well participants can detect changes in stimulation, a modified center-out task was performed. As shown in Figure 5c, the three starting points for this task were labeled by their corner in the stimulation parameter space. The first point corresponded with the lowest intensity and slowest frequency (Bottom-Left). The second point corresponded to the highest intensity but the slowest flutter frequency (Top-Left), and the last point corresponded to the fastest flutter frequency and lowest intensity (Bottom-Right). In each trial of a task, stimulation began at one of the three starting points and stayed constant for one second. From there, the stimulation parameters changed in one of three ways: only the intensity, only the flutter frequency, or both intensity and flutter frequency simultaneously. The parameters changed by ± 30% at a constant rate over 1.5 seconds, with the direction of change being tied to the starting point tested and staying there for an added second. Participants moved the joystick along one of the three directions and pressed a button once they had reached their final position. The number of times a target was correctly identified was recorded as a correct percentage. At the end of the performance of all three extremes-in tasks, a verbal survey was performed in which participants were asked which of the three tasks they preferred.

### Study Protocol

The study protocol consisted of a calibration, after which participants performed the center-out tasks starting with the intensity-only version followed by the flutter frequency only. After these, either all three versions of the discrete control task or all three versions of the extremes-in task were performed in a row. The order of the versions was chosen at random. All participants received training for each task just prior to data collection.

### Statistical Analysis

Statistical analysis was conducted using the SciPy library in Python 3 and the R software. The dataset was checked for normality using a Shapiro-Wilks test. One tailed T-tests or Mann-Whitney tests were used to compare each target’s performance to chance. The chance level depends on the number of targets presented to the participant. Chance performance was set at 33%, 25%, and 20% in tasks with three, four, and five targets, respectively. ANOVAs were used to assess inter-task differences. Unless otherwise specified, the significance level was set to α = 0.05. If a post-hoc test was called for, a Tukey’s post-hoc test was performed. Chi-square analyses were used to evaluate whether the overall performance of a task was better than chance. Data is presented as mean ± standard deviation.

## Data Availability

The data that support the findings of this study are openly available at the following:

https://doi.org/10.5061/dryad.1rn8pk12q

## Acknowledgments

This work was supported by a Department of Defense US Army Joint Warfighter Medical Research Program (JWMRP) grant award (W81XWH1910839). TB was also partly supported by an I^3^R Graduate Research Assistantship. I would like to thank Dr. Samantha Robinson for her guidance with the statistical analysis.

## Data Availability Statement

The data that support the findings of this study are openly available at the following: https://doi.org/10.5061/dryad.1rn8pk12q

